# An AI Method for Assessing Coding Consistency in a Large Dataset

**DOI:** 10.1101/2024.01.17.24301268

**Authors:** Stuart J. Nelson, Ying Yin, Yijun Shao, Phillip Ma, Mark S. Tuttle, Qing Zeng-Treitler

## Abstract

**Objective:** We developed a method to assess the consistency of the assignment of ICD codes, using coding performed at a United States health system at the time of the transition from ICD-9CM to ICD-10CM.

**Methods:** Using clusters of equivalent codes derived from the US Centers for Disease Control General Equivalence Mapping (GEM) tables, ICD assignments occurring during the ICD-9CM to ICD-10CM transition were evaluated in EHR data from the US Veterans Administration Central Data Warehouse, using a deep learning model based on 860 covariates. The model was then used to detect abrupt changes across the transition; additionally changes at each VA station were examined.

**Results:** Many of the 687 most-used code clusters had ICD-10CM assignments differing greatly from that predicted by the GEM from the codes used in ICD-9CM. Notably, the observed transition patterns varied widely across care locations.

**Conclusion:** Machine learning can model variability across time and across location, enabling an assessment of coding consistency. Expert review is not scalable, deep learning model applied to a large dataset of EHR records provides an approximation of ground truth.

## Introduction

The adoption of Electronic Health Record (EHR) systems was an effort to improve patient care by improving the accuracy and clarity of medical records and by increasing the availability of the record to patients and physicians. Additionally, EHR data supports the study of patient outcomes in real-world settings. As a result EHR data has become widely used for observational clinical research to improve post-care surveillance, and to perform comparative effectiveness studies.[1] However, the usefulness of EHR-based research depends centrally on the quality of the data to be found there.[2]

Traditionally, EHR data quality is measured along 5 dimensions: 1. completeness (e.g., the amount of missing data), 2. correctness (e.g. a plausible distribution of values), 3. concordance (e.g., the use of terminology standards), 4. plausibility (e.g., out of range values), and 5. currency (e.g., the timeliness or recency of data).[3] None of these measures looks specifically at the quality of coding.

There are challenges when transforming raw EHR data into a dataset for analysis.[4] One such challenge is the need to define a cohort, often to ascertain the presence or absence of a disease or phenotype. Most often this is accomplished by using standard codes, particularly the International Classification of Diseases (ICD). Even as researchers have begun using other data (e.g. using free-text notes through natural language processing (NLP)) to augment phenotype definitions,[5-7] the ICD codes remain a critical piece in defining cohorts for study.[8-11] While we might expect codes to be used consistently over time and across different healthcare organizations and facilities, the degree of consistency has not been established.

The U.S. transition from coding using ICD, 9th Revision, Clinical Modification (ICD-9CM) to ICD, 10th Revision, Clinical Modification (ICD-10CM) in the United States, provided an opportunity to conduct a formal investigation of coding consistency in a large EHR database. The difficult transition in coding from using ICD-9CM (with approximately 14,000 diagnostic codes) to using ICD-10CM (with about 70,000 codes, providing for greater specificity in the codes) was eased somewhat by the General Equivalence Mapping (GEM) tables provided by the Centers for Disease Control (CDC). Despite those efforts, however, marked changes in the frequency of certain conditions were observed by some researchers.[12-15] Accordingly, we developed a method for systematic investigation into the consistency (and therefore the accuracy) of ICD coding, using the transition as an exemplar.

#### Statement of Significance

##### Problem

Data quality measures do not address inconsistency in coding. Inconsistencies affect cohort and phenotype creation.

##### What is Known

There were marked changes in the frequency of equivalent codes assigned after the transition from ICD-9CM to ICD-10CM.

##### What This Paper Adds

A method was developed using machine learning to detect aberrancies in coding. Wide variation in coding was found at the time of transition. Wide variation in the variance was found between locations.

## Methods

We used the United States (US) Veteran Affairs (VA) Corporate Data Warehouse (CDW), with 20 million patients and with up to 20 years of follow-up, 4 billion outpatient visits, and 16 million inpatient visits (numbers as of Aug 2022). The US VA health care system has 1,298 health care facilities, including 171 VA Medical Centers and 1,113 outpatient sites. Because of its size and diverse locations, it provided and excellent testbed.

### GEM Mapping

We identified clusters of codes made up from both versions of ICD that were suggested to be equivalent by the GEM. The GEM tables are not symmetric; we ignored the mapping directions and merged the mapped pairs of codes to yield a single combined GEM table; it included four types of ICD-9CM to ICD-10CM code mappings: one-to-one, one-to-many, many-to-one, and many-to-many.

We constructed new diagnosis variables (clusters) with GEM mappings (version 2018) as building blocks. If the whole GEM table is a bipartite graph, in which nodes are codes and edges are correspondence relations between the two versions of codes, then the GEM mappings are exactly the connected components (as defined in graph theory). These mappings connected all the components of the two versions. The sets of codes in the fully connected subgraphs are then referred to as clusters. Each code is present in one, and only one, cluster; a total of 6,707 clusters were identified.

The frequency of use of the ICD-9/10 CM codes within one year of the ICD 9/10 transition (between 10/1/2014 and 9/30/2016) was used to identify the number of patients with each diagnostic code. Limiting the codes for further study to those assigned to at least 0.1% of the observed population resulted in 700 clusters.

### Preparing the Data for Investigation

The frequency with which the clusters were used at each station of the VA on a weekly basis was obtained by summing the frequency of the codes in the cluster. The weekly frequency of use of each cluster was summed across the entire VA system.

The data was prepared for multivariate time-series analysis. The weekly visit counts of each cluster at each station were collected from 2001 to 2019, a total of 992 weeks and 2.6 billion visits. Candidate observables included: 1) total weekly inpatient and outpatient visit counts; 2) total patient counts by gender, race, and ethnicity; 3) top 500 weekly lab test counts grouped by analyte group; and 4) weekly drug fill counts grouped by VA drug class (based on pharmacological effects). Because the visit count has seasonal patterns, the number of the week in the year was also added. This resulted in a total of 860 observables to be used as predictors of the ICD code cluster. We expected to address “over-fitting” concerns through the large number of observations.

### Deep Learning Model

A two-branch deep learning model was designed for time series forecasting. The first branch of the model processing temporal data was composed of a linear projection layer, a positional encoding layer, and 2 Transformer blocks resembling those in the original Transformer architecture for natural language processing. For a given week t, the prior N weekly data points X _t-N+1_, …, X _t_ were used as the input data. Each data point X_t_ was a scaled vector containing both weekly visit count for a given cluster and covariate values. The linear projection layer then projected each data point to a vector of dimension d_model_. The positional encoding layer mapped the corresponding position of each data point in the sequence to a vector of dimension d_model_ using sine and cosine functions. The positions were numbered as 1 through N starting from X_t_ and backward. The element-wise sum of the two sequences of vectors were then fed into the 2 Transformer blocks.

The second branch, processing the input data containing the covariate information of week t+1 was a feedforward neural network (FFNN) with residual connection. The outputs of the two branches were then combined by element-wise addition. The final output went through another FFNN with residual connection before reaching the output layer. The output layer had only one node with no nonlinear activation functions and output the predicted (re-scaled) visit count for the week t+1. The loss function for training was the mean squared error function.

For each cluster, a model was trained separately. N=12 weekly data points predicted the next future week visit count. The stochastic gradient descent with Nesterov momentum method was used as the optimizer. The mean square error of the predicted and the observed counts was calculated for model evaluation. The dropout method was used for regularization.

### Applying the Predictions of the Modes

The fitted model was then applied to the fiscal years 2016-2019 to predict the usage of the cluster-equivalent ICD-10CM codes. We focused on the period between 10/1/2015 and 9/30/2016 to detect significant changes at the time of the transition. We computed the difference between the predicted value and the actual value of the frequency of use. The standard deviation of the residuals in the training phase (1/1/2001 - 9/30/2015) provided a measure of the variability of the difference between the prediction and the actual values. We calculated the Absolute value of the Standardized Residual (ASR) (the residual divided by the standard deviation) to measure the significance of changes. For each cluster mode, two sets of ASR values were computed: ASR_1,_ the ASR of the first week after transition (10/4/2015) and ASR_2_, the averaged ASR of the year after transition. This enabled us to measure both the immediate abrupt change and year trend change. Aberrance was defined as an ASR value greater than 4.

Using each station as its own control and allowing each station to be its own control (to account for possible variations in prevalence of diseases) we calculated the rate of change of each cluster frequency at the time of the transition from ICD-9CM to ICD-10CM.

## Results

Dramatic code use changes were observed in many clusters after the ICD transition (Figure 1).

**Figure 1:**
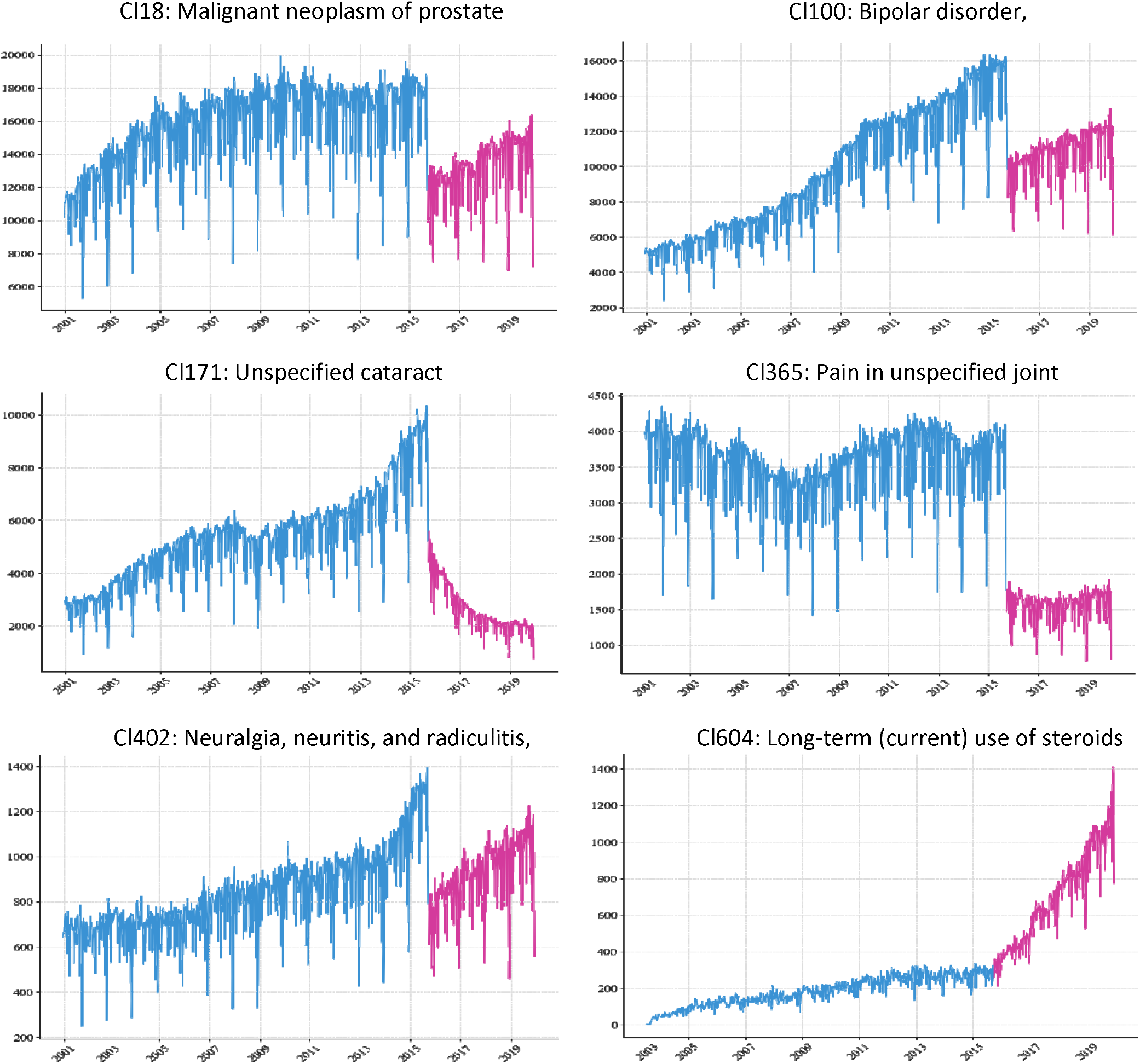
Examples of weekly visit trend of diagnosis clusters.

We noted wide variance between differing VA stations in the way the codes were used (Figure 2).

**Figure 2:**
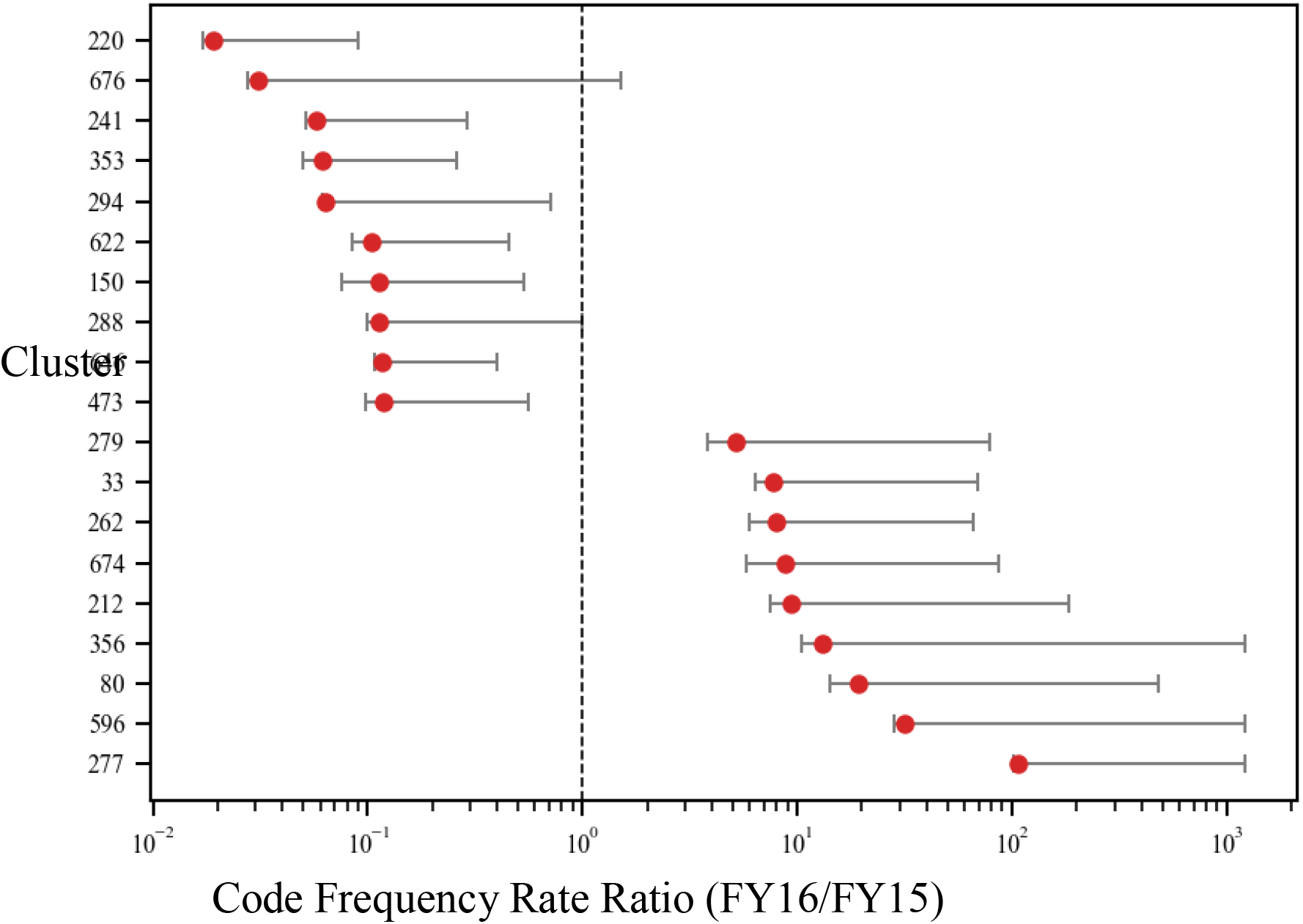
**Variation of code frequency rate changes among VA stations of top 20 most changed clusters. The whisker range cover 95% of all variations and excludes outliers**.

Of the 700 clusters studied, 13 clusters were mapped with ICD-10CM codes effective after 2017 and thus no data was available for the testing phase. Of the remaining 687 clusters, 323 (47.0%) exhibited a significant abrupt change with ASR_1_ exceeding 4.

Expanding the analysis to the year after transition, the median ASR_2_ of the 687 clusters was 3.94 for the DL model indicating the code usages of more than half of the clusters underwent dramatic shifts in the year after transition, and the aberrant signals were not just limited to one week. To confirm the findings, using human review of the 98 clusters for changes at the time of transition as the reference standard, we obtained area under the curve (AUC) value of 93.6% for the DL model.

## Discussion

**Interesting questions arise when these results are viewed. The first question is, of course, “why?” What could lead to such discrepancies in coding behavior? Did the GEM tables fail to provide any usable guidance in coding for established patients?**

Our analysis found that the use of the codes in the clusters formed from the GEM deviated significantly from what was predicted. A large proportion of the clusters showed a deviance of 4 standard deviations or more in their predicted use. Are there data aberrancies not attributable to the flaws in GEM?

The diversity in the patterns of the changes in code use between VA stations is curious. Bearing in mind that the prevalence of diseases may be variable depending on location, using each station as its own control, and looking at the differences in the rate of change allows the elimination of prevalence as a cause for the diversity. While this finding may not be surprising, the extent of diversity quantifies what may have been only suspected.

**There is another question which should concern all who use coded data in observational studies. How can changes occurring during a transition be managed? Cimino [16] provided an example of what happened with changing (and reuse) of codes in ICD, and the difficulty was appreciated [17]. While this study of something of largely historical interest, with the anticipated changeover to ICD 11 our findings should lead reviewers to consider what will happen during that transition**.

**The implications of inconsistencies in coding lead to recognition that very careful attention must be paid to defining cohorts and phenotypes for all their uses. Are the methods used to define these cohorts and phenotypes adequate to overcome the difficulties inherent in the inconsistencies occurring at different times and different locations?**

**We propose this deep learning method as one way to find an approximation of ground truth. When trying to assess reliability of findings (such as in an estimate of AUC), it is necessary to provide such an estimation of ground truth. While expert review is generally considered an acceptable means, providing expert review for large data sets may be cumbersome and not feasible. Recently another approach recognized the lack of scalability in defining a cohort or phenotype. It also used structured data and proposed a framework for knowledge representation of that data. [18]**.

With the increasing acceptance of natural language processing (NLP), some may feel that structured data quality issue has become less important. While our team has been active in the development of NLP-based phenotypes, we believe that NLP complements structured data but cannot replace it. One example is the Phenotype Knowledgebase (PheKB) which contains dozens of phenotypes sourced from eMerge and other projects. About half of these phenotypes involved NLP while almost all of them involved ICD codes and other structured data. We also note that in some large databases (e.g., Cerner Real World Data (now known as Oracle Life Sciences), IBM Market Scan, and NIH N3C) only structured data is available, precluding the ability to use any NLP techniques.

## Conclusions

This investigation suggests that a serious data quality issues in large EHR datasets can be detected using machine learning, particularly so-called deep learning. Few studies check the longitudinal use of structured data codes for consistency. We believe more efforts need to be invested to monitor and improve the quality of EHR data for health services and clinical research.

## Data Availability

The data is available through the US Veterans Administration.

## Acknowledgements

This work was supported by VA HSRD grant 1I21HX003278-01A1, and by AHRQ grant R01 HS28450-01A1.

